# Convergent cross-sectional and longitudinal evidence for gaming-cue specific posterior parietal dysregulations in early stages of Internet Gaming Disorder

**DOI:** 10.1101/2020.01.18.20018036

**Authors:** Fangwen Yu, Rayna Sariyska, Bernd Lachmann, Qianqian Wang, Martin Reuter, Bernd Weber, Peter Trautner, Shuxia Yao, Christian Montag, Benjamin Becker

## Abstract

Exaggerated reactivity to drug-cues and emotional dysregulations represent key symptoms of early stages of substance use disorders. The diagnostic criteria for (Internet) Gaming Disorder strongly resemble symptoms for substance-related addictions. However, previous cross-sections studies revealed inconsistent results with respect to neural cue reactivity and emotional dysregulations in these populations. To this end the present fMRI study applied a combined cross-sectional and prospective longitudinal design in excessive online gamers (n=37) and gaming-naïve controls (n=67). To separate gaming-associated changes from predisposing factors, gaming-naive subjects were randomly assigned to 6 weeks of daily Internet gaming or a non-gaming condition. At baseline and after the training subjects underwent an fMRI paradigm presenting gaming-related cues and non-gaming related emotional stimuli. Cross-sectional comparisons revealed gaming-cue specific enhanced valence attribution and neural reactivity in a parietal network, including the posterior cingulate/precuneus in excessive gamers as compared to gaming naïve-controls. Prospective analysis revealed that six weeks of gaming elevated valence ratings as well as neural cue-reactivity in a similar parietal network, specifically the posterior cingulate/precuneus in previously gaming-naïve controls. Together, the prospective longitudinal design did not reveal supporting evidence for altered emotional processing of non-gaming associated stimuli in excessive gamers while convergent evidence for increased emotional and neural reactivity to gaming-associated stimuli was observed. Findings suggest that exaggerated neural reactivity in posterior parietal regions engaged in self-referential processing already occur during early stages of regular gaming probably promoting continued engagement in gaming behavior.

## Introduction

In May 2019 the World Health Organization (WHO) included *Gaming Disorder* as distinct diagnosis (6C51) in the category *disorders due to addictive behaviors* in the International Classification of Diseases (ICD, 11^th^ version). Previously, the American Psychiatric Association (APA) included *Internet Gaming Disorder* as an emerging disorder in the appendix of DSM-5.^1^ Diagnostic criteria strongly resemble those for substance-related addictions and include (1) compulsive gaming and lack of control over gaming, (2) preoccupation with gaming at the expense of other activities, and (3) continued gaming despite negative consequences (see also [2]).

The inclusion in the diagnostic classification systems takes account of the growing concerns with respect to detrimental mental health effects of excessive and in some cases compulsive engagement in Internet gaming. Prevalence estimates of Internet Gaming Disorder (IGD) range from 0.3%-27.5% worldwide^3^ with a recent representative survey suggesting that 1.16% of adolescents in Germany meet the diagnostic criteria for IGD according to DSM-5 criteria^4^ (prevalence estimates according to the WHO criteria see [5]). Accumulating evidence from cross-sectional and longitudinal surveys suggest detrimental effects on mental health which partly resemble those observed in substance addiction, with elevated emotional distress, anxiety and depression being among the most commonly reported.^4, 6^

In line with the symptomatic overlap between substance addictions and IGD an increasing number of cross-sectional studies combined functional MRI with validated experimental paradigms from substance addiction research to determine IGD-associated neural alterations in the domains of cognition and reward-related processing, including exposure to gaming associated cues (cue reactivity). An early meta-analysis encompassing cognitive and reward-related fMRI studies reported that - compared to healthy controls – excessive gamers exhibited increased neural activation in fronto-cingulate regions, including anterior as well as posterior cingulate cortices (ACC, PCC) during cognitive and reward-related paradigms.^7^ A subsequent meta-analysis^8^ that incorporated a broader range of reward-related and executive functions confirmed generally exaggerated reactivity in fronto-cingulate circuits and additionally demonstrated increased reactivity in fronto-striatal circuits, particularly dorsal striatal and lateral prefrontal regions, with some evidence for domain-specific alterations in the latter regions depending on the motivational context of the cognitive domains assessed. Two recent meta-analyses specifically focused on cue-reactivity in IGD and reported increased neural reactivity in lateral prefrontal, cingulate and dorsal striatal regions in response to gaming cues within IGD individuals^9^, and exaggerated cue-reactivity in lateral prefrontal and posterior parietal regions, including the PPC and precuneus, as well as decreased insular activation in IGD individuals relative to controls.^10^

Notably, in contrast to convergent evidence for exaggerated ventral striatal reactivity in response to drug-associated cues across substance addicted populations^11, 12^ the ventral striatum was not found to be robustly engaged in IGD. The ventral striatum is strongly engaged in signaling reward expectancy, reinforcement behavior and salience with convergent translational evidence suggesting that this region critically contributes to the initial development of addiction via mediating reinforcing effects of the drug as well as associated incentive salience and learning processes whereas the dorsal striatum mediates habitual and compulsive use during later stages of the disorder.^13, 14^ In line with animal models ventral striatal cue reactivity has already been observed in regular, non-dependent, drug users whereas dependent users during later stages of the disorder exhibit pronounced reactivity in the dorsal striatum engaged in habit formation.^15-17^

A recent framework by Brand et al. extended previous models on substance-related addictions to internet-related addictive behaviors, including gaming, and emphasized the importance to account for interactions of person-affect-cognition and execution variables (I-PACE).^18^ A recent update of this model emphasized the relevance to (1) consider predisposing vulnerability factors such as emotional dysregulations, e.g. anxiety or deficient reward processing, as well as (2) to differentiate early and later stages in addiction development, suggesting that the initial stages of behavioral addictions are mediated by the ventral striatum dependent incentive urges and sensitization with progressive impairments in prefrontal stimulus-specific inhibitory control during later stages of the disorder.^19^

However, in contrast to accumulating evidence for altered emotional processing in regular and dependent substance users emotional dysregulations in IGD have not been examined.^20-22^ Furthermore, based on extensive translational animal models^14, 23^ the important role of ventral striatal cue reactivity during early as well as later stages of substance use disorders has been established^15, 24, 25^, whereas previous studies on cue-reactivity in IGD revealed inconsistent findings and due to the cross-sectional retrospective design remain limited with respect to determining cue-associated changes during the early stages of excessive, regular gaming.

To this end the present study applied a combined cross-sectional prospective longitudinal design in n = 45 excessive gamers of the massively multiplayer online role-playing game (MMORPG) World of Warcraft (WOW) and n = 83 gaming-naïve controls. Following the baseline (T1) assessment non-gaming controls were randomly assigned to 6 weeks of daily WOW gaming (training group, TRG) or no gaming (training control group, TCG) (detailed protocols see also [26]). An MMORPG was employed due to the high popularity and particular high risk of escalation of use of these games^27, 28^. At both timepoints subjects underwent fMRI while they were presented WOW-associated as well as non-gaming associated positive and negative visual stimuli to asses both neural cue reactivity and emotional processing. To additionally examine alterations on the level of subjective experience behavioral ratings of valence and arousal were assessed following the fMRI assessments. Based on the previous literature we expected (1) altered emotional processing in gamers versus non-gamers at baseline reflecting predisposing alterations or changes related to regular excessive gaming, and (2) exaggerated gaming cue-reactivity in fronto-striatal and fronto-cingulate regions in excessive gamers relative to controls, as well as increased fronto-striatal (particularly ventral striatal) cue reactivity in formerly gaming naïve participants following the training.

## Materials and methods

### Participants

The study was part of a larger cross-sectional and prospective project examining the effects of excessive Internet gaming on brain structure and function. The sample was identical to our previous study examining effects of internet gaming on brain structure.^26^ *N*=131 healthy participants (71 males, 60 females; mean age = 23.80, SD = 3.97) were enrolled in the project and underwent MRI assessments twice, at T1 (baseline) and after an interval of six weeks (T2). All participants were free of a history of, or current psychiatric/neurological disorders, drug abuse or apparent brain structural abnormalities. Participants in the WoW gaming group (WoW group) had to fulfill the following inclusion criteria: (1) WoW gaming > 7 hours a week (years of WoW gaming 6.28, SD = 2.10) (2) play other multiplayer game < 7 hours a week; and, 3) no first-person shooter gaming to control for potential adaptive changes in prefrontal emotion regulation during exposure to negative emotional stimuli.^29^ Participants in the control group were WoW gaming naïve at enrollment and reported generally low online video gaming addiction tendencies (**table 1**). Based on the study exclusion criteria and MRI brain structural quality assessments n =12 subjects were excluded (details see [26]). From the remaining 119 subjects additional n = 15 subjects were excluded from the present study due to excessive head motion or technical issues during functional fMRI (details see flow diagram presented in supplement materials **Figure S1**) leading to a final sample size of *N*=104 participants (54 males and 50 females, mean age = 23.51, SD = 3.91; WOW, n = 37; control group, n = 34; training group, n = 33). For avoiding duplicate analysis with the previous study^26^ and consistency within the image analysis, questionnaire data assessing levels of Internet addiction and behavior data during fMRI were analyzed from present sample (n = 104). To determine cue-reactivity and emotional alterations associated with excessive WOW gaming cross-sectional and prospective analyses were applied. For the cross-sectional comparison data from the initial assessment (T1) of the WoW group (n = 37; 23 males and 14 females, mean age = 24.84, SD = 4.27) were compared to the non-gaming group (n = 67; 31 males and 36 females, mean age = 22.78, SD = 3.52). To determine alterations in a prospective longitudinal fashion the non-gaming group was randomly assigned to two different six weeks interventions following T1 data acquisition: the training group was required to play WoW at least 1 hour per day (TRG, n = 33), while the training control group (TCG, n = 34) did not engage in the game. All participants provided written informed consent and received monetary compensation. Addiction severity was assessed using previously validated self-report scales. WOW gaming addiction in the gamers was assessed using the WOW gaming addiction scale.^30^ General online video gaming addiction (OVGA) at T1 and training associated changes were assessed using a modified version of the version of the video game addiction questionnaire^31^ which was administered to all subjects at both timepoints (T1, T2). For details on the gaming addiction assessments see also [26].

**Table 1.** Group characteristics: Demographics and Internet Gaming.

| Sample(n=104) | TCG<br>M(SD) | TRG<br>M(SD) | WoW gamer<br>M(SD) | F($\chi^2/t$) | p |
| --- | --- | --- | --- | --- | --- |
| Age(years) | 22.4(3.00) | 23.2(3.99) | 24.8(4.27) | 3.93 | 0.023* |
| Gender(male/female) | 18/16 | 13/20 | 23/14 | 3.61 | 0.165 |
| OVGA T1 | 7.85(1.86) | 7.52(1.48) | 15.10(5.23) | 55.40 | <0.001*** |
| OVGA T2 | 8.09(3.10) | 8.85(2.65) | 14.80(4.79) | 35.00 | <0.001*** |
| WoW addiction T1 | - | - | 86.70(24.00) | - | - |
| WoW addiction_T2 | - | 53.80(17.20) | 82.90(21.90) | - | - |

OVGA\_T1: measured on-line computer game addiction scores at T1; OVGA\_T2: measured on-line computer game addiction scores at T2; WoW addiction\_T1: measured WoW addiction questionnaire at T1; WoW addiction\_T2: measured WoW addiction questionnaire at T2.
\* $p < 0.05$ , \*\* $p < 0.01$ , \*\*\* $p < 0.001$

The study and procedures were in accordance with the latest Declaration of Helsinki and had full ethical approval of the local ethic committee at the University of Bonn, Bonn, Germany.

### Experimental paradigm

Cue-reactivity and emotional processing was assessed using a modified version of a fMRI block-design paradigm that has been previously demonstrated to be sensitive to substance dependence and gaming associated neural changes.^15, 20, 29^. During the paradigm visual stimuli from four categories were presented: negative, positive, neutral and WoW pictures. Matched arousal and valence ratings for negative and positive pictures were derived from the International Affective Picture system (IAPS) database, while WoW pictures were screenshots from the game. During fMRI the stimuli were presented in condition-specific blocks with six blocks per condition each encompassing 5 stimuli presented for 4 seconds. The blocks were presented in a pseudorandomized order while ensuring that each block was followed by a block of a different category. To ensure attentive processing all of the blocks were followed by the presentation of an additional picture following a 1.5s (on average, randomized between 1000ms and 2000ms) black screen and subjects had to indicate via button press whether the picture had been shown during the preceding block (see supplement materials **Figure S2**). Blocks were separated by a fixation cross period (randomized between 3000ms and 6000ms) during which a fixation cross was displayed that served as low level baseline. Total duration of the paradigm was approximately 12 minutes.

### Behavioral assessments – emotional processing

To further assess differences in subjective emotional experience participants were asked to rate arousal and valence of stimuli following the MRI acquisition. For each participant the rating pictures were randomly selected from all pictures in the paradigm. Rating was conducted by displaying rating scales ranging from 1-7 (indicating: arousal, 1 very low – 7 very high, valence, 1 very negative – 7 very positive).

### MRI data acquisition

Functional MRI data was acquired using the following acquisition parameters: [T2*-weighted echo-planar images (EPI)] was acquired on a 1.5 T Siemens Magnetom Avanto Siemens Scanner (Siemens, Erlangen, Germany), using 12 channel standard matrix head coil [31 axial slices; 3 mm slice-thickness; 3 ⨯ 3 ⨯ 3.3 mm voxel size; TR was 2.5 seconds; TE was 45 milliseconds; FoV = 192 mm ⨯ 192 mm; Flip angle = 90°; matrix size = 64 ⨯ 64; axial orientation, applied filter prescan normalization, PAT modus GRAPPA 2 with 32 reference lines]. T1-weighted structural image was acquired before T2*-weighted image and used to improve normalization of the functional images (acquisition parameters: 160 sagittal slices; TR = 1.660 seconds; TE = 3.09 ms; Flip angle 15°; FoV = 256 ⨯ 256, matrix size = 256 ⨯ 256; 1 ⨯ 1 ⨯ 1 mm resolution; sagittally oriented 3D sequence, magnetization preparation non-selective inversion recovery with TI = 850 ms). The paradigm was presented using Presentation software (https://www.neurobs.com/index_html) in combination with an in-house developed template for paradigm programming.

### MRI data preprocessing

Data was preprocessed using the DPABI toolbox^32^ (version V4.3_171210, http://rfmri.org/DPARSF). The first 10 functional volumes were discarded to allow for MRI equilibration. Preprocessing included standardized preprocessing procedures including realignment to correct for head motion, co-registration of the mean functional image to the brain structural image and normalization using a two-step procedure including segmentation of the structural image and subsequent application of the corresponding normalization parameters to the functional time series (Montreal Neurological Institute standard space, MNI, resampled at 3 ⨯ 3 ⨯ 3mm). To account for inverse consistent deformations in image registration a fast diffeomorphic registration algorithm (Diffeomorphic Anatomical Registration using Exponentiated Lie Algebra, DARTEL^33^) was applied for both, segmentation and smoothing. The normalized images were smoothed using a full width at half maximum (FWHM) Gaussian filter with 6mm.

### fMRI data analysis

To determine effects of WOW gaming on cue reactivity and emotional processing, two-level random effects general linear model (GLM) analyses was conducted in SPM12b (https://www.fil.ion.ucl.ac.uk/spm) On the 1^st^ level the GLM was applied to model the condition-specific blocks and the attention test phase following each block. To further control for motion related artifacts the six head-movement were included in the design matrix as additional regressors. The first level matrix was convolved with the standard hemodynamic response function (HRF). Contrasts of interest from the 1^st^ level analyses were subjected to second level voxel-wise ANOVA models as implemented in SPM12b.

In line with the cross-sectional prospective design of the study two second level analyses were conducted. To examine cue-reactivity and emotion processing in excessive gamers a voxel-wise whole brain mixed ANOVA analyses including group as between-subject factor (WoW vs. non-gamer) and condition as between-subject factor (Neg, Neu, Pos, WoW) was calculated on the baseline (T1) data. Second, to determine cue-reactivity changes during the early stages of gaming a voxel-wise whole brain mixed ANOVA was conducted including training group (TCG vs. TRG) as between subject factor and timepoint (T1 vs. T2) as within subject factor. To further disentangle significant interaction effects parameter estimates were extracted from the corresponding regions from 6 mm radius spheres centered at the peak coordinates using marsbar 0.44 (http://marsbar.sourceforge.net/).

## Results

### Demographics and gaming behavior

Age and gender were examined as potential confounders (see **table 1**). In the present sample (n = 104), there was no differences in gender distribution between the three groups (*χ(2)*^2^ = 3.61, *p* = 0.165). However, WoW gamers were slightly older that than TCG (mean difference [MD] _WoW-TCG_ = 2.5, t_101_ = 2.75, *p*_Bonferroni_ = 0.021), whereas there was no age difference between TCR and TRG (MD_TRG-TCG_ = 0.9, t_101_ = 0.92, *p*_Bonferroni_ = 1.000), or WoW group and TRG (MD_WoW-TRG_ = 1.6, t_101_ = 1.78, *p*_Bonferroni_ = 0.232). This pattern was consistent with the larger sample reported in our previous study (for details see [26]) and consequently age was controlled for in analyses including comparisons of the WOW group with the gaming-naïve groups.

### Addiction severity – cross sectional comparison at time point 1 (T1)

A one-way ANOVA of online video game addiction (OVGA) scores at T1 (see **table 1**) (n = 104) including three groups (TCG, TRG, WoW) as between-group factor revealed a significant interaction among three groups (F_(2)_ = 55.40, p < 0.01, *η*^2^ =0.526). The post-hoc tests revealed that the WoW group had higher OVGA scores than both, TCG (MD_(WoW-TCG)_ = 7.21, t_(100)_ = 8.82, *p*_Bonferroni_ < 0.001) and TRG (MD_(WoW-TRG)_ = 7.54, t_(106)_ = 9.23, *p*_Bonferroni_ < 0.001) at baseline assessment. There was no difference between TCG and TRG (MD_(TRG-TCG)_ = 0.33, t_(100)_ = 0.4, *p*_Bonferroni_ = 1.000). These results were consistent with the original sample (details see [26]).

### Cross-sectional analysis – subjective emotional experience

Cross-sectional examination of valence ratings (T1) by means of a mixed ANOVA revealed a significant interaction effect between groups (non-gamer and WoW gamers) and emotion (Neg, Neu, Pos, WoW; F _(3,255)_ = 8.88, *p* < 0.001, *η*^2^ = 0.014) with post hoc analyses demonstrating specifically higher valence ratings for WOW-associated stimuli in the gamers (MD_(WoW gamer - non-gamer)_ = 0.770, t_(55)_ = 4.72, *p*_*Bonferroni*_ < 0.001) yet no differences in the other conditions (p > 0.05, **Fig 1a**). For arousal ratings a mixed ANOVA revealed no significant interaction effects between the factor group and the factor of emotion category (F _(3,255)_ = 1.09, *p* =0.352, *η*^2^ = 0.003, **Fig 1 b**). Findings remained stable after including age as covariate in the analyses (details see supplement materials, **tables S1-S3**).

**Figure 1.**
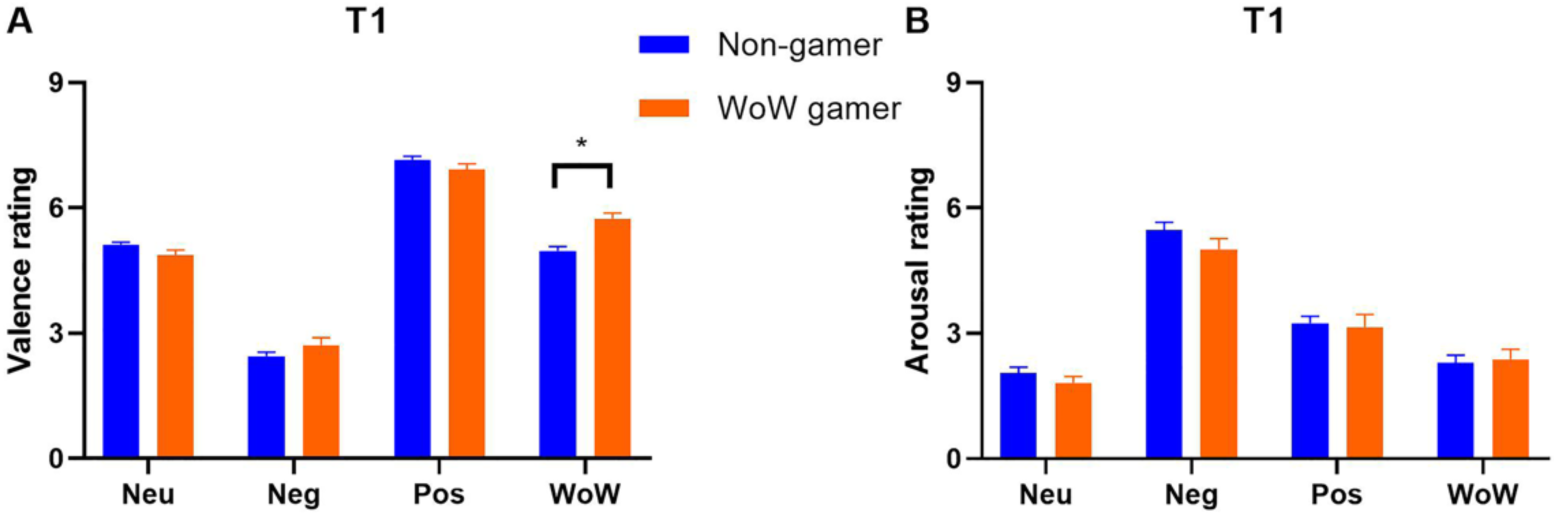
Cross-sectional analysis of subjective emotional experience in terms of valence and arousal. (A) Excessive gamers reported higher positive valence for the WOW related stimuli as compared to the non-gamers; (B) With respect to arousal no significant group differences were observed. Abbreviations: WoW, World of Warcraft, Neu, neutral stimuli, Neg, negative stimuli; Pos, positive stimuli; WoW, World of Warcraft (gaming) stimuli; T1, timepoint 1 (baseline) assessment

### Cross-sectional analysis – functional MRI

In line with the analyses of the behavioral data a mixed ANOVA with the factor groups (Non-gamer vs. WoW gamer) as between-subject factors and emotional conditions (Neg, Pos, Neu and WoW) as within-subject factors was computed. On the whole brain level, a significant interaction effect was observed in four clusters located in the posterior cingulate cortex (PCC), right precuneus (rPrecuneus), right inferior parietal lobule (rIPL), and left inferior parietal lobule (lIPL) (p < 0.05 FDR-corrected, see **table 2** and **Figure 2A**). Post hoc analyses that directly compared the two groups on the condition-specific extracted parameter estimates revealed that excessive gamers demonstrated exaggerated reactivity in these regions in response to WOW stimuli, whereas no neural alterations during processing of emotional stimuli were observed (all p-values > 0.05 after Bonferroni multiple comparison correction, **Figure 2B**). These results were stable after controlling for age (details see supplement materials **tables S4-S7**). Given that the analyses of the behavioral and neural cross-sectional data convergently revealed specific differences between gamers and non-gamers in response to WOW-related stimuli the prospective longitudinal analysis of the training effects focused on the WOW condition.

**Table 2.**
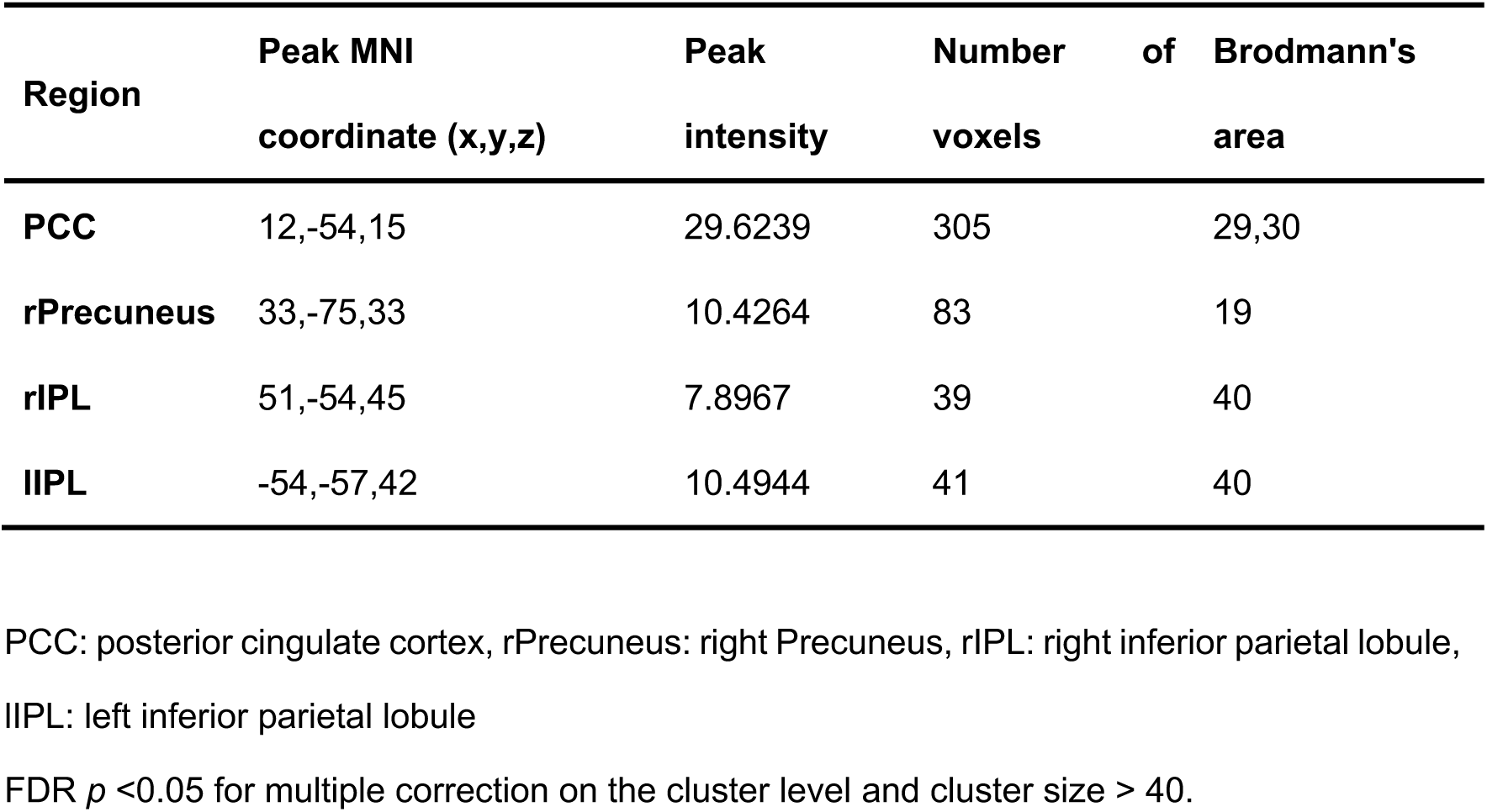
Group differences in brain activation - cross-sectional analysis.

**Figure 2.**
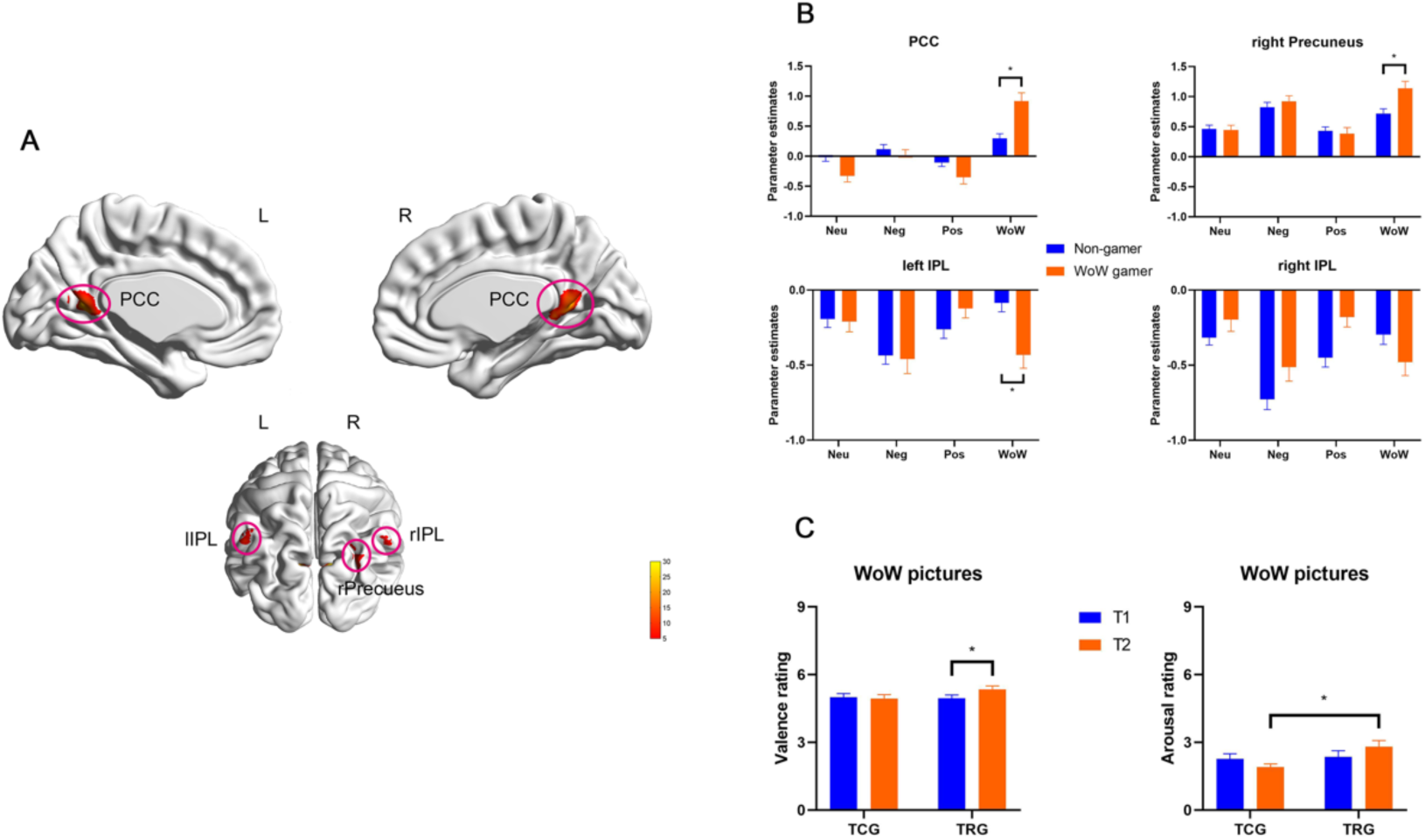
Cross-sectional analysis of the fMRI paradigm including positive, negative, neutral and WOW gaming related stimuli. (A) Voxel-wise mixed ANOVA model including group and emotional condition revealed a significant interaction effect in four clusters located in the PCC, lIPL, rIPL, rPrecuneus (FDR *p* < 0.05, cluster size k > 39, color bar represents peak intensity); (B) Post hoc analysis revealed that excessive gamers exhibited higher neural cue-reactivity in response to gaming-related pictures compared to non-gamer (Bonferroni-corrected *p* < 0.05); (C) Results from the longitudinal analysis of emotional experience ratings. Results from post hoc tests demonstrating that the training group (TRG) reported increased valence ratings after the training relative to the baseline ratings, and higher arousal ratings after the training as compared to the non-training (TCG) control group at T2. Abbreviations: TCG, training control group; TRG, training group; WoW, World of Warcraft, Neu, neutral stimuli, Neg, negative stimuli; Pos, positive stimuli; WoW, World of Warcraft (gaming) stimuli; T1, timepoint 1 (baseline) assessment; PCC: posterior cingulate cortex, IPL: inferior parietal lobule, lIPL: left inferior parietal lobe, rIPL: right inferior parietal lobe, rPrecuneus: right precuneus * *p* < 0.05, ** *p* <0.01, *** *p* < 0.001.

### Longitudinal analysis of online gaming addiction changes during training

For the longitudinal analysis of OVGA, we conducted a 2 x 3 mixed ANOVA including timepoints (T1 vs. T2) as within subject factor and groups (TCG, TRG, WoW gamer) as between subject factor in the present sample (n=104). Result showed a significant interaction effect between timepoints and groups (F_(2,99)_ = 3.56, *p* = 0.032, *η*^2^ = 0.006) with the TRG demonstrating a trend for an increase in OVGA scores over the six-week training (MD_TRG(T2-T1)_ = 1.33, t _(99)_ =2.80, *p* _Bonferroni_ = 0.093). The post hoc test showed that the WoW group had higher OVGA scores than TCG and TRG both at T1 and T2 (T1: MD_(WoW gamer –TCG)_ = 7.13, t _(133)_ =8.39, *p* _Bonferroni_ < 0.001; MD_(WoW gamer –TRG)_ = 6.37, t _(133)_ =7.50, *p* _Bonferroni_ < 0.001; T2: MD_(WoW gamer –TCG)_ = 6.73, t _(133)_ =7.90, *p* _Bonferroni_ < 0.001; MD_(WoW gamer –TRG)_ = 5.96, t _(133)_ =7.01, *p* _Bonferroni_ < 0.001), and there is no difference between TCG and TRG both at T1 and T2 (see **table 1**, *p* > 0.05 after Bonferroni correction). These results were consistent with data of the original sample presented in Zhou et al. (2019).

### Prospective longitudinal analyses – effects of training on subjective emotional experience

Mixed ANOVAs that focused on the training effects on the emotional reactivity to gaming associated cues by including the training groups as between subject factor (TRG, TCG) and timepoints (T1, T2) as within subject factor revealed a significant interaction effect (F_(1,55)_ = 6.26, *p* = 0.015, *η*^2^ = 0.017) for valence ratings with post hoc analysis suggesting that valence ratings significantly increased in the TRG (MD_(T2-T1)_ = 0.395, t_(55)_ = 1.75, *p*_*Bonferroni*_ = 0.022, **Fig 2C**) but not in the TCG. An additional ANOVA on the arousal ratings also revealed a significant group by timepoint interaction effect (F _(1,55)_ = 6.36, *p* = 0.015, *η*^2^ = 0.025), with post hoc analysis indicating that the training group reported higher arousal ratings after the training as compared to the TCG for WOW stimuli (MD_(TRG-TCG)_ = 0.91, t_(85.9)_ = 2.73, *p*_*Bonferroni*_ = 0.046, **Fig 2C**).

Together with the increase in the OVGA scores the changes in the subjective perception of the WOW stimuli indicate that the training intervention was efficient in terms of inducing an initial tendency for addictive gaming behavior/habit formation.

### Prospective longitudinal analyses – effects of training on neural cue reactivity

Examination of training-related changes by means of a voxel-wise whole brain ANOVA including the between-subject factor group (TRG vs. TCG) and the within-subject factor timepoint (T1 vs. T2) for game-related pictures (WoW pictures) revealed significantly larger changes in the PCC, rPrecuneus, left middle occipital gyrus (lmOG) and lIPL (*p* < 0.05, FDR corrected) in the training group (see **table 3** and **Fig 3A**). To determine the overlap between changes associated with long-term excessive WOW gaming (cross sectional analysis) and short-term WOW-training the networks from both analysis approaches were overlaid. Results revealed common alterations in two regions encompassing the PCC and rPrecuneus (**table 4** and **Fig 3B**). Subsequent extraction of parameter estimates from these regions revealed that WOW-related neural reactivity increased in both regions in the training relative to the control group over the course of the six weeks intervention period (two sample t-test, for PCC, MD _(TRG-TCG)_ = 0.72, SE _(MD)_ = 0.22, t _(65)_ = 3.33, *p* = 0.001; for rPrecuneus, MD _(TRG-TCG)_ = 0.66, SE _(MD)_ = 0.16, t _(65)_ = 3.33, *p* < 0.001; **Fig 3C**). Additional control analysis on the condition-specific extracted parameter estimates from these regions revealed no changes in the PCC and precuneus reactivity to the non-gaming related stimuli (neutral, positive, negative; all *p*_*bonferroni*_ > 0.05) arguing against unspecific training-induced changes in these regions.

**Table 3.**
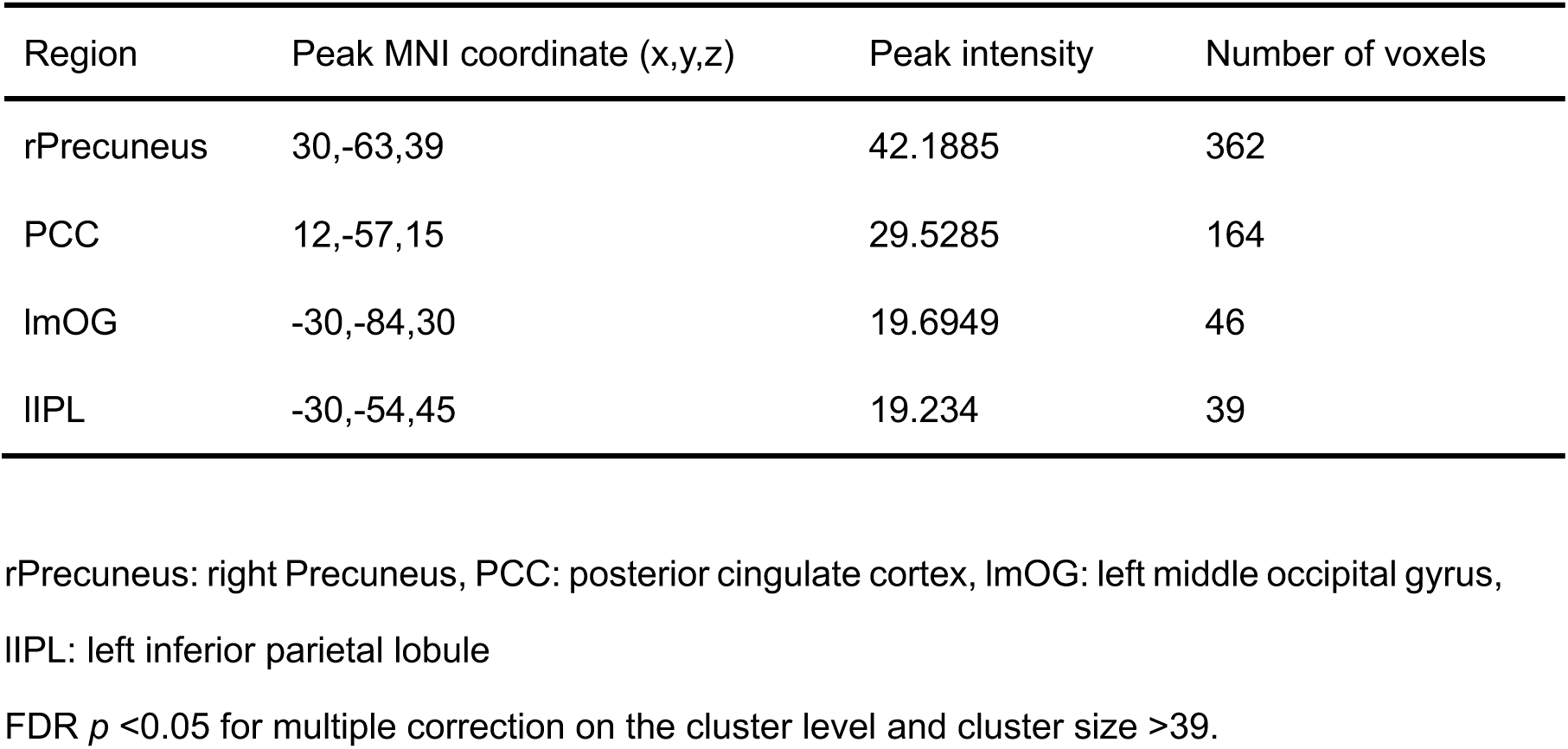
Group differences in brain activation – prospective analysis.

**Table 4.**
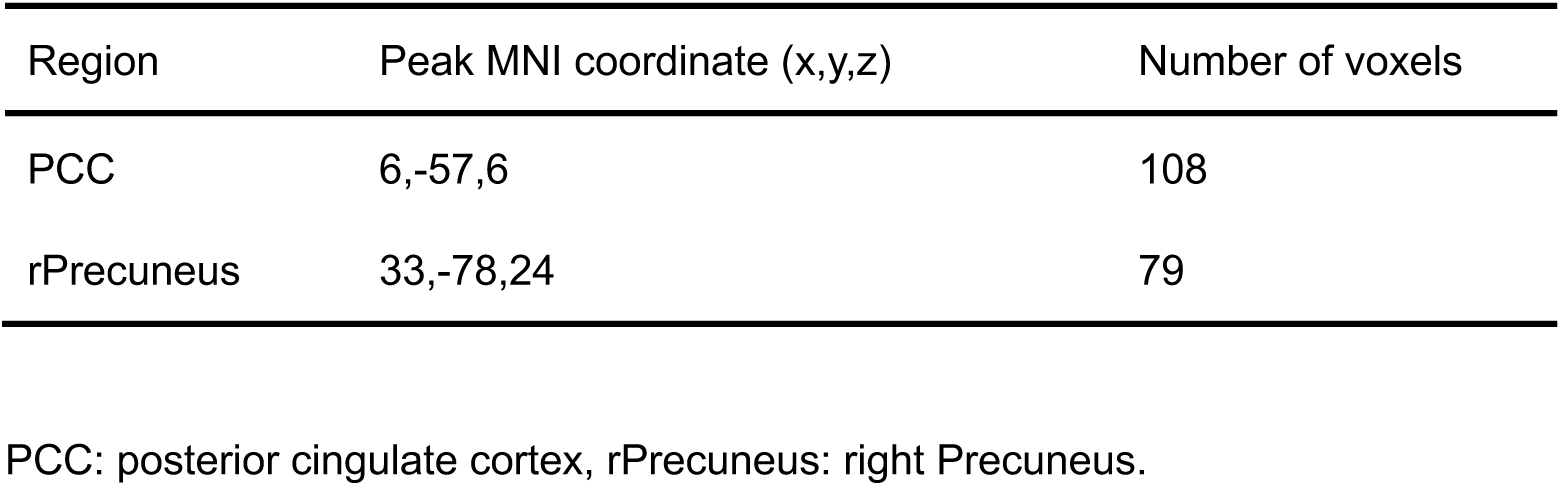
Overlap between brain functional alterations in the cross-sectional and prospective analysis.

| Region | Peak MNI coordinate (x,y,z) | Number of voxels |
| --- | --- | --- |
| PCC | 6,-57,6 | 108 |
| rPrecuneus | 33,-78,24 | 79 |

PCC: posterior cingulate cortex, rPrecuneus: right Precuneus.

**Figure 3.**
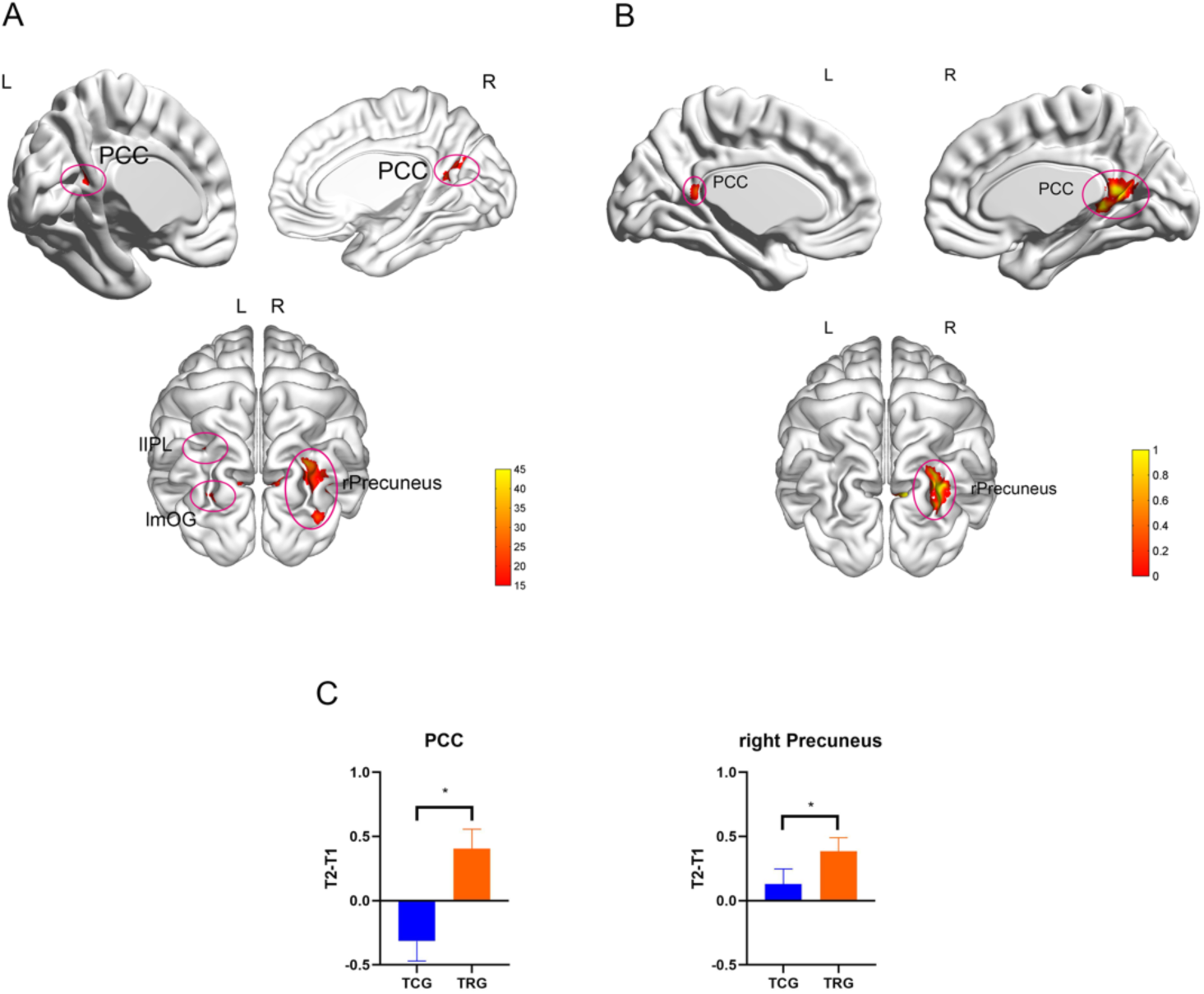
Prospective longitudinal analysis of gaming effects on neural cue-reactivity. (A) Voxel-wise mixed ANOVA model including group and timepoint revealed an interaction effect in four clusters located in the PCC, rPrecuneus, lIPL, and lmOG (FDR *p* < 0.05, cluster size k > 39, color bar represents peak intensity) (B) Overlaying changes observed in the cross-sectional and prospective analysis revealed that the PCC and rPrecuneus demonstrated convergent alterations across the analyses; (C) Post hoc examination of change scores (T2>T1) in these regions revealed that the training group exhibited significantly increased reactivity to the gaming related stimuli after the training (*p* < 0.05) Abbreviations: TCG, training control group; TRG, training group; WoW, World of Warcraft; PCC, posterior cingulate cortex; rPrecuneus, right Precuneus; IPL, left inferior parietal lobe; lmOG, eft middle occipital gyrus * *p* < 0.05

### Examination of neural cue-reactivity within the excessive gamers

In contrast to our expectations and some previous studies, including one study using a comparable WOW sample and cue-reactivity paradigm^34^ neither the cross sectional nor the longitudinal analysis revealed cue-induced striatal activation. To further delineate the cue-reactivity networks and the robustness of this response we explored neural reactivity within the group of excessive WOW gamers (WOW > neutral) separately at T1 and T2. At both time-points the WOW cues elicited robust activation in a highly similar network engaging the default mode network (DMN) including the PCC and mPFC, cognitive control network (CCN) including MFG and IFG, and posterior parietal attention network including precuneus and IPL, whereas no reactivity in striatal regions was observed (**Fig. 4**, details presented in **Table S8**).

**Figure 4.**
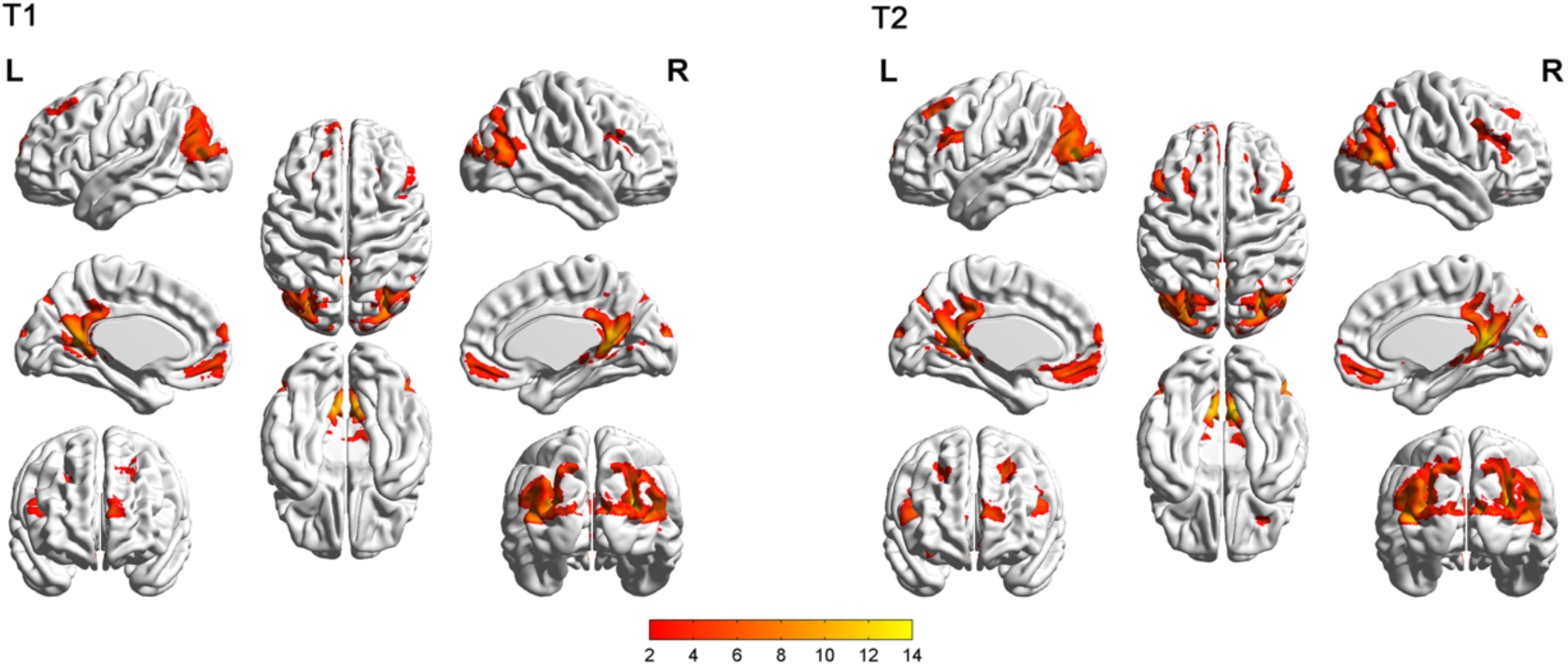
Examination of neural cue-reactivity (WoW pictures > Neutral pictures) within the excessive gamers at both timepoints (T1 and T2). At both time-points, the gaming cues elicited robust activation in the bilateral posteriot cingulate cortex (PCC), ventral medial prefrontal cortex (vmPFC), inferior prefrontal gyrus (IFG), middle prefrontal gyrus (MFG), superior prefrontal gyrus (SFG), parietal lobe, middle temporal gyrus (MTG), occipital lobe (details see also supplemental materials table S12).

## Discussion

The present fMRI study used a combined cross-sectional prospective longitudinal design to examine cue-reactivity and emotional processing during early stages of Internet Gaming Disorder. Cross-sectional comparisons revealed enhanced positive valence attribution and neural reactivity in a parietal network, including the PCC/precuneus and the bilateral IPL, in excessive gamers as compared to gaming naïve-controls. Alterations in excessive gamers were specifically observed during processing of gaming-related stimuli whereas the gamers exhibited normal processing of non-gaming related emotional stimuli. The prospective analyses revealed that six weeks of gaming increased valence ratings as well as neural cue-reactivity in a similar parietal network, specifically the PCC/precuneus, in previously gaming-naïve controls. Finally, further examination of gaming cue-reactivity within the excessive gamers revealed robust neural cue-reactivity at both timepoints in a fronto-parietal network encompassing the posterior (PCC/precuneus) and anterior (medial PFC) DMN, core regions of the frontal cognitive control network (CCN) and the posterior parietal attention network, including the IPL, whereas no neural cue-reactivity in striatal regions was observed.

In line with previous studies in behavioral addiction^9^, including IGD^8, 10^ the gaming-related stimuli induced stronger behavioral and neural responses in excessive gamers as compared to gaming-naïve controls. In contrast to accumulating evidence for emotional alterations, particularly elevated negative affect and depressive symptom load^5, 35^, in excessive gamers, as well as an increasing number of studies reporting altered behavioral and neural processing of non-drug associated emotional stimuli in individuals with regular drug use (e.g [21]), excessive gamers in the present study did not demonstrate altered emotional processing of non-gaming related stimuli. A previous cross-sectional study reported altered fronto-striatal processing of negative stimuli in IGD individuals^36^, suggesting that IGD partly resembles emotion-regulation deficits as previously observed in regular substance users.^20^ Participants in the previous IGD study fulfilled the DSM-5 criteria for IGD and compared to the excessive gamers in the present study may therefore represent a sample that already made the transition from excessive to problematic and finally compulsive gaming. In the context of the previous findings the present results may reflect that (1) in contrast to regular substance users excessive gamers present intact emotional processing, (2) predisposing emotional alterations may render excessive gamers vulnerable for the transition to IGD, or alternatively that (3) only during later stages of IGD emotional processes become progressively dysregulated.

In line with recent meta-analytic data suggesting exaggerated neural responses to gaming cues in IGD, excessive gamers exhibited enhanced neural reactivity in response to gaming related pictures, in both cross-sectional comparison with gaming-naïve controls as well as relative to neutral pictures. Exaggerated reactivity to drug or behavioral addiction related cues has been consistently reported in the literature and documented across substance addictions^11, 12^, pathological gambling^37^ and increasingly IGD.^8, 10^ The exaggerated neural reactivity to addiction-associated cues in these populations is considered to reflect an acquired dopamine-mediated reinforcement-based learning process that develops with repeated pairing of the stimuli with rewarding experience. Neural cue-reactivity thus reflects mechanisms related to the development of exaggerated reward and salience attribution as well as self-directed processes and habit formation. Neural cue reactivity accordingly engages several brain systems engaged in these processes, specifically the striatum engaged in reward processes and habit formation, frontal regions engaged in executive control as well as parietal regions engaged in attention and self-referential processes.^38^

In substance-based addictions neural cue-reactivity has been most consistently observed in striatal regions involved in reward processes and habit formation.^11, 12, 38^ Ventral striatal cue reactivity has been already observed in non-addicted, regular substance users which is considered to reflect exaggerated incentive salience of drug-associated cues.^15, 17^ In contrast to some previous studies in IGD subjects^39^, the present study did not find robust evidence for exaggerated striatal cue-reactivity in excessive gamers, suggesting that alterations in striatal reward processing regions may only occur during later stages of the disorder. On the other hand, neural cue reactivity in posterior parietal regions was observed in the cross-sectional analysis as well as in the prospective longitudinal analysis. The observation that six weeks of gaming increased valence perception as well as posterior parietal cue-reactivity in previously gaming-naïve individuals suggests that changes in this region are critically related to regular (daily) engagement in gaming and already occur following relatively short time-periods. Specifically, the PCC and precuneus demonstrated convergently increased reactivity towards gaming related cues. Both regions represent core hubs of the posterior DMN engaged in internally oriented attention and self-referential processing^40^ and have been repeatedly reported in cue-reactivity studies in substance addiction^15, 41^ as well as in meta-analytic studies on cue-reactivity in IGD (e.g. [10]) and have been suggested to reflect a stronger engagement of self-referential processes.^42^ Moreover, as core region of the DMN the PCC/precuneus not only promote internally guided self-relevant processes but also the detection of self-relevant information in the environment (e.g. [43]). Based on these observations a recent study employed transcranial stimulation (TMS) to this region during cue-induced reactivity in substance addiction and demonstrated that down-regulation of this region normalized exaggerated self-referential attribution in cannabis-dependent subjects.^44^ In the context of these previous findings the convergent cross sectional and prospective findings on exaggerated neural cue reactivity in the PCC/precuneus may point to an important role of self-referential processes in the initial stages of excessive gaming.

Together, the present combined prospective longitudinal design did not reveal supporting evidence for behavioral or neural alterations during the processing of *non-gaming* associated stimuli in (excessive) gamers while convergent evidence for increased emotional and neural reactivity to gaming-associated stimuli was observed. Findings suggest that exaggerated neural reactivity in posterior parietal regions engaged in self-referential processing already occurs during early stages of regular gaming probably promoting continued engagement in gaming behavior.

## Data Availability

Data is available from the authors upon request.

## Acknowledgements

This work was supported by the National Key Research and Development Program of China (Grant No. 2018YFA0701400 to B.B.); Fundamental Research Funds for Central Universities (ZYGX2015Z002 to B.B.); and Science, Innovation and Technology Department of the Sichuan Province (2018JY0001 to B.B.). C.M. is funded by a Heisenberg Grant awarded to him by the German Research Foundation (DFG) (MO 2363/3-2). The authors declare no conflict of interest.

## Authors Contribution

CM designed the study. RS, BL and PT contributed to the acquisition of the data. FY, BB and CM analyzed the data, interpreted the results and drafted the manuscript. MR, BW, QW and SY provided critical revision of the manuscript for important intellectual content.

## References

1. Petry, N.M. and C.P. O’Brien, Internet gaming disorder and the DSM-5. Addiction, 2013. 108(7): p. 1186–1187.

2. Pontes, H.M., et al., Measurement and conceptualization of Gaming Disorder according to the World Health Organization framework: The development of the Gaming Disorder Test. International Journal of Mental Health and Addiction, 2019: p. 1–21.

3. Mihara, S. and S. Higuchi, Cross-sectional and longitudinal epidemiological studies of I nternet gaming disorder: A systematic review of the literature. Psychiatry and clinical neurosciences, 2017. 71(7): p. 425–444.

4. Rehbein, F., et al., Prevalence of Internet gaming disorder in German adolescents: Diagnostic contribution of the nine DSM-5 criteria in a state-wide representative sample. Addiction, 2015. 110(5): p. 842–851.

5. Montag, C., et al., Psychopathological Symptoms and Gaming Motives in Disordered Gaming— A Psychometric Comparison between the WHO and APA Diagnostic Frameworks. Journal of clinical medicine, 2019. 8(10): p. 1691.

6. Krossbakken, E., et al., A cross-lagged study of developmental trajectories of video game engagement, addiction, and mental health. Frontiers in psychology, 2018. 9: p. 2239.

7. Meng, Y., et al., The prefrontal dysfunction in individuals with Internet gaming disorder: a meta-analysis of functional magnetic resonance imaging studies. Addiction biology, 2015. 20(4): p. 799–808.

8. Yao, Y.-W., et al., Functional and structural neural alterations in Internet gaming disorder: A systematic review and meta-analysis. Neuroscience & Biobehavioral Reviews, 2017. 83: p. 313–324.

9. Starcke, K., et al., Cue-reactivity in behavioral addictions: A meta-analysis and methodological considerations. Journal of behavioral addictions, 2018. 7(2): p. 227–238.

10. Zheng, H., et al., Meta-analyses of the functional neural alterations in subjects with Internet gaming disorder: Similarities and differences across different paradigms. Progress in Neuro-Psychopharmacology and Biological Psychiatry, 2019: p. 109656.

11. Chase, H.W., et al., The neural basis of drug stimulus processing and craving: an activation likelihood estimation meta-analysis. Biological psychiatry, 2011. 70(8): p. 785–793.

12. Kühn, S. and J. Gallinat, Common biology of craving across legal and illegal drugs–a quantitative meta - analysis of cue - reactivity brain response. European Journal of Neuroscience, 2011. 33(7): p. 1318–1326.

13. Di Chiara, G. and A. Imperato, Drugs abused by humans preferentially increase synaptic dopamine concentrations in the mesolimbic system of freely moving rats. Proceedings of the National Academy of Sciences, 1988. 85(14): p. 5274–5278.

14. Everitt, B.J. and T.W. Robbins, Drug addiction: updating actions to habits to compulsions ten years on. Annual review of psychology, 2016. 67: p. 23–50.

15. Zhou, X., et al., Cue Reactivity in the Ventral Striatum Characterizes Heavy Cannabis Use, Whereas Reactivity in the Dorsal Striatum Mediates Dependent Use. Biological Psychiatry: Cognitive Neuroscience and Neuroimaging, 2019.

16. Brumback, T., et al., Adolescent heavy drinkers’ amplified brain responses to alcohol cues decrease over one month of abstinence. Addictive behaviors, 2015. 46: p. 45–52.

17. Vollstädt-Klein, S., et al., Initial, habitual and compulsive alcohol use is characterized by a shift of cue processing from ventral to dorsal striatum. Addiction, 2010. 105(10): p. 1741–1749.

18. Brand, M., et al., Integrating psychological and neurobiological considerations regarding the development and maintenance of specific Internet-use disorders: An Interaction of Person-Affect-Cognition-Execution (I-PACE) model. Neuroscience & Biobehavioral Reviews, 2016. 71: p. 252–266.

19. Brand, M., et al., The Interaction of Person-Affect-Cognition-Execution (I-PACE) model for addictive behaviors: update, generalization to addictive behaviors beyond internet-use disorders, and specification of the process character of addictive behaviors. Neuroscience & Biobehavioral Reviews, 2019.

20. Zimmermann, K., et al., Emotion regulation deficits in regular marijuana users. Human brain mapping, 2017. 38(8): p. 4270–4279.

21. Zimmermann, K., et al., Altered orbitofrontal activity and dorsal striatal connectivity during emotion processing in dependent marijuana users after 28 days of abstinence. Psychopharmacology, 2018. 235(3): p. 849–859.

22. Charlet, K., et al., Neural activation during processing of aversive faces predicts treatment outcome in alcoholism. Addiction biology, 2014. 19(3): p. 439–451.

23. Markou, A., et al., Animal models of drug craving. Psychopharmacology, 1993. 112(2-3): p. 163–182.

24. MacNiven, K.H., et al., Association of Neural Responses to Drug Cues With Subsequent Relapse to Stimulant Use. JAMA network open, 2018. 1(8): p. e186466-e186466.

25. Guillem, K. and S.H. Ahmed, Incubation of Accumbal Neuronal Reactivity to Cocaine Cues During Abstinence Predicts Individual Vulnerability to Relapse. Neuropsychopharmacology, 2018. 43(5): p. 1059.

26. Zhou, F., et al., Orbitofrontal gray matter deficits as marker of Internet gaming disorder: converging evidence from a cross-sectional and prospective longitudinal design. Addiction biology, 2019. 24(1): p. 100–109.

27. Nagygyörgy, K., et al., Typology and sociodemographic characteristics of massively multiplayer online game players. International journal of human-computer interaction, 2013. 29(3): p. 192–200.

28. Smyth, J.M., Beyond self-selection in video game play: An experimental examination of the consequences of massively multiplayer online role-playing game play. CyberPsychology & Behavior, 2007. 10(5): p. 717–721.

29. Montag, C., et al., Does excessive play of violent first-person-shooter-video-games dampen brain activity in response to emotional stimuli?Biological psychology, 2012. 89(1): p. 107–111.

30. Peters, C.S. and L.A. Malesky Jr, Problematic usage among highly-engaged players of massively multiplayer online role playing games. CyberPsychology & Behavior, 2008. 11(4): p. 481–484.

31. Lemmens, J.S., P.M. Valkenburg, and J. Peter, Development and validation of a game addiction scale for adolescents. Media psychology, 2009. 12(1): p. 77–95.

32. Yan, C. and Y. Zang, DPARSF: a MATLAB toolbox for” pipeline” data analysis of resting-state fMRI. Frontiers in systems neuroscience, 2010. 4: p. 13.

33. Ashburner, J., A fast diffeomorphic image registration algorithm. Neuroimage, 2007. 38(1): p. 95–113.

34. Sun, Y., et al., Brain fMRI study of crave induced by cue pictures in online game addicts (male adolescents). Behavioural brain research, 2012. 233(2): p. 563–576.

35. Bonnaire, C. and D. Baptista, Internet gaming disorder in male and female young adults: The role of alexithymia, depression, anxiety and gaming type. Psychiatry research, 2019. 272: p. 521–530.

36. Yip, S.W., et al., Is neural processing of negative stimuli altered in addiction independent of drug effects? Findings from drug-naïve youth with internet gaming disorder. Neuropsychopharmacology, 2018. 43(6): p. 1364.

37. Clark, L., I. Boileau, and M. Zack, Neuroimaging of reward mechanisms in Gambling disorder: an integrative review. Molecular psychiatry, 2019. 24(5): p. 674–693.

38. Zilverstand, A., et al., Neuroimaging impaired response inhibition and salience attribution in human drug addiction: a systematic review. Neuron, 2018. 98(5): p. 886–903.

39. Liu, L., et al., Activation of the ventral and dorsal striatum during cue reactivity in Internet gaming disorder. Addiction biology, 2017. 22(3): p. 791–801.

40. Andrews-Hanna, J.R., et al., Functional-anatomic fractionation of the brain’s default network. Neuron, 2010. 65(4): p. 550–562.

41. Filbey, F.M., et al., Marijuana craving in the brain. Proceedings of the National Academy of Sciences, 2009. 106(31): p. 13016–13021.

42. Zilverstand, A., M.A. Parvaz, and R.Z. Goldstein, Neuroimaging cognitive reappraisal in clinical populations to define neural targets for enhancing emotion regulation. A systematic review. Neuroimage, 2017. 151: p. 105–116.

43. Luber, B., et al., Self-enhancement processing in the default network: a single-pulse TMS study. Experimental brain research, 2012. 223(2): p. 177–187.

44. Prashad, S., et al., Testing the role of the posterior cingulate cortex in processing salient stimuli in cannabis users: an rTMS study. European Journal of Neuroscience, 2019. 50(3): p. 2357–2369.

